# Randomized trials on non-pharmaceutical interventions for COVID-19 as of August 2021: a meta-epidemiological analysis

**DOI:** 10.1101/2021.08.20.21261687

**Authors:** Julian Hirt, Perrine Janiaud, Lars G. Hemkens

## Abstract

**Background:** Numerous non-pharmaceutical interventions (NPIs) were taken worldwide to contain the spread of the COVID-19 pandemic. We aimed at providing an overview of randomized trials assessing NPIs to prevent COVID-19.

**Methods:** We included all randomized trials assessing NPIs to prevent COVID-19 in any country and setting registered in ClinicalTrials.gov and the World Health Organization International Clinical Trials Registry Platform using the COVID-evidence platform (until 17 August 2021). We searched for corresponding publications in MEDLINE/PubMed, Google Scholar, the Living Overview of Evidence platform (L-OVE), and the Cochrane COVID-19 registry as well as for results posted in registries.

**Results:** We identified 41 randomized trials. Of them, 11 were completed (26.8%) including 7 with published results. The 41 trials planned to recruit a median of 1,700 participants (IQR, 588 to 9,500, range 30 to 35,256,399) with a median planned duration of 8 months (IQR, 3 to 14, range 1 to 24). Most came from the United States (n=11, 26.8%). The trials mostly assessed protective equipment (n=11, 26.8%), COVID-19-related information and education programs (n=9, 22.0%), access to mass events under specific safety measures (n=5, 12.2%), testing and screening strategies (n=5, 12.2%), and hygiene management (n=5, 12.2%).

**Conclusions:** Worldwide, 41 randomized trials assessing NPIs have been initiated with published results available to inform policy decisions for only 7 of them. A long-term research agenda including behavioral, environmental, social, and systems level interventions is urgently needed to guide policies and practices in the current and future public health emergencies.

## Introduction

Numerous non-pharmaceutical interventions (NPIs) were taken worldwide to contain the COVID-19 pandemic.^1^ NPIs are considered crucial to prevent infections, in particular for non-vaccinated populations.^2^ Such interventions during COVID-19 include drastic social distancing and lockdown measures impacting billions of people. The need for randomized evidence on NPIs has been highlighted on numerous occasions ^3-5^. However, no randomized trials assessing NPIs were initiated early in the pandemic ^6^ and it is likely their number have remained sparse. ^7-9^ We aimed to provide a systematic overview of the research agenda of randomized trials assessing NPIs to prevent COVID-19.

## Methods

A meta-epidemiological analysis was performed to collect the current trials on NPIs and to summarize their status and characteristics. We did not register this study or published a specific study protocol. We adapted PRISMA 2020 to provide a comprehensive study report, where applicable.^10^

### Eligibility criteria

This meta-epidemiological analysis included all registered randomized trials assessing NPIs to prevent COVID-19 in any country and setting. We considered any NPIs as single measures or in combination involving, for example, information and education interventions, protective equipment, disinfection, social distancing, isolation, quarantine, testing, or community-wide containment strategies. We excluded interventions based on drugs, biologicals, vaccines, herbals, traditional medicine, and homeopathy. Trials assessing NPIs without any health outcome related to SARS-CoV-2-infections were excluded (e.g., interventions aiming to improve vaccination rates).

### Search and selection of trials

We searched the COVID-evidence platform (www.covid-evidence.org) for eligible trials as of 17 August 2021. COVID-evidence is a freely available continuously updated database that contains information about worldwide planned, ongoing, and completed randomized trials on any intervention to treat or prevent SARS-CoV-2-infections. Trials registered in ClinicalTrials.gov and the World Health Organization International Clinical Trials Registry Platform (ICTRP) are retrieved on a weekly basis. COVID-evidence uses a multimethod approach combining peer-reviewed search strategies of study registries, continuous automated extraction of search results, automated classifications combined with manual screening and data extraction (details on the methodology are published elsewhere ^11^). For all NPI eligible trials identified in COVID-evidence, we searched for corresponding publications (peer-reviewed and preprints) in MEDLINE/PubMed, Google Scholar, the Living Overview of Evidence platform (L-OVE), and the Cochrane COVID-19 registry using trial registration numbers (until 19 August 2021). We also recorded if trial results were reported in the registries. All searches and the selection of trials were done by one researcher (JH or PJ). Unclear cases were discussed and resolved within the study team (JH, PJ, LGH).

### Data extraction

One researcher (JH) extracted data on trial status, start to end date, country, setting, number of enrolled individuals and clusters, topic (details on NPIs and characteristics of the population), and design features (randomization unit, number of arms, outcome measurement). Extractions were tabulated and verified by two other researchers (PJ, LGH).

### Data analysis

Descriptive statistics using numbers and percentages were used. We used R (version 4.1).

## Results

We identified 41 eligible randomized trials (Figure; Table). As of August 2021, 11 trials were completed (26.8%) including 7 with published results, 17 ongoing (41.5%), 11 not yet recruiting (26.8%), and 2 terminated early or withdrawn (5.9%). Most came from the United States (n=11, 26.8%), United Kingdom (n=4, 9.8%), and France (n=4, 9.8%). Of note, none of the completed and terminated trials had made their results available on the registries.

**Figure.**
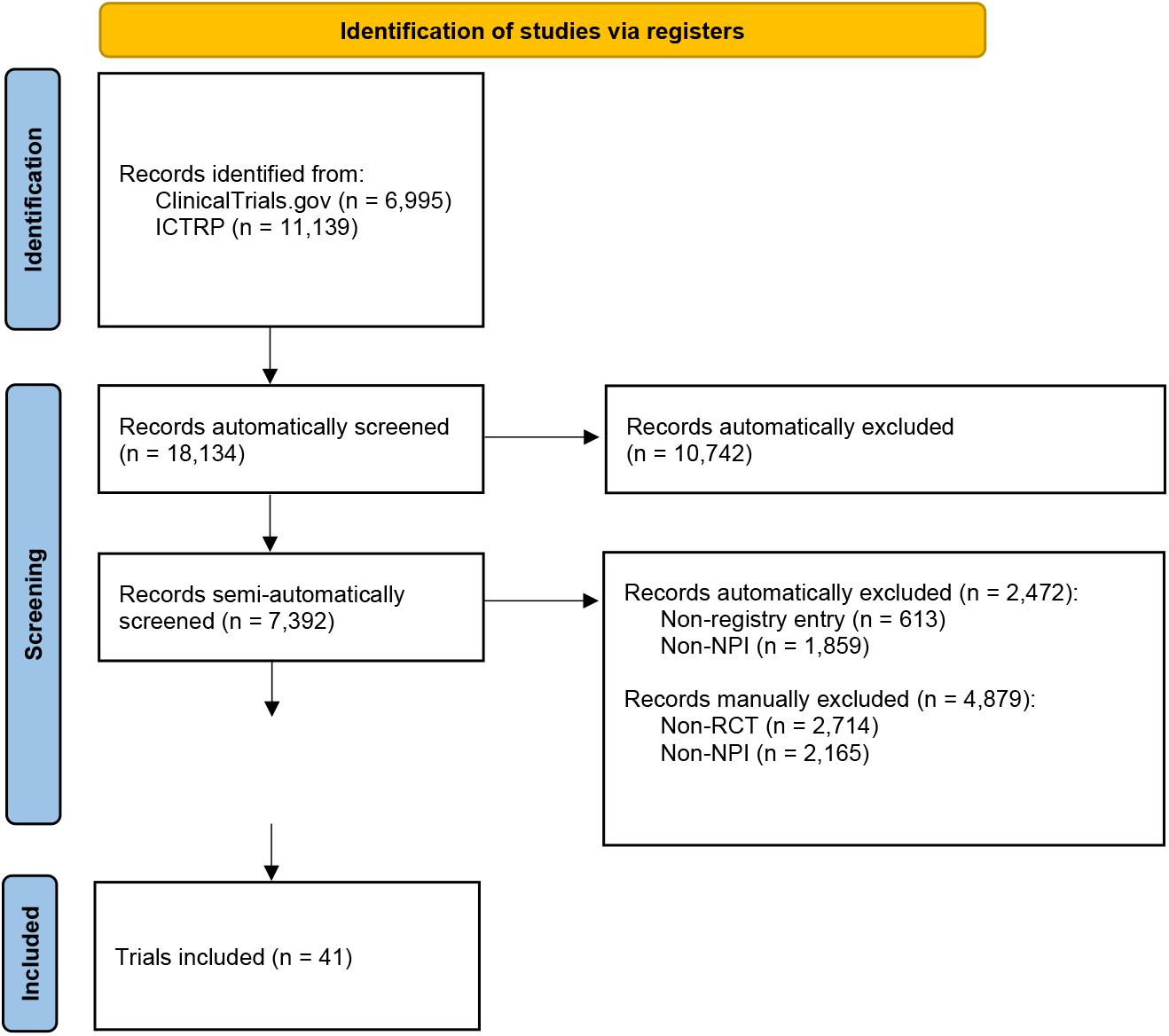
Retrieval and trial selection process

**Table.**
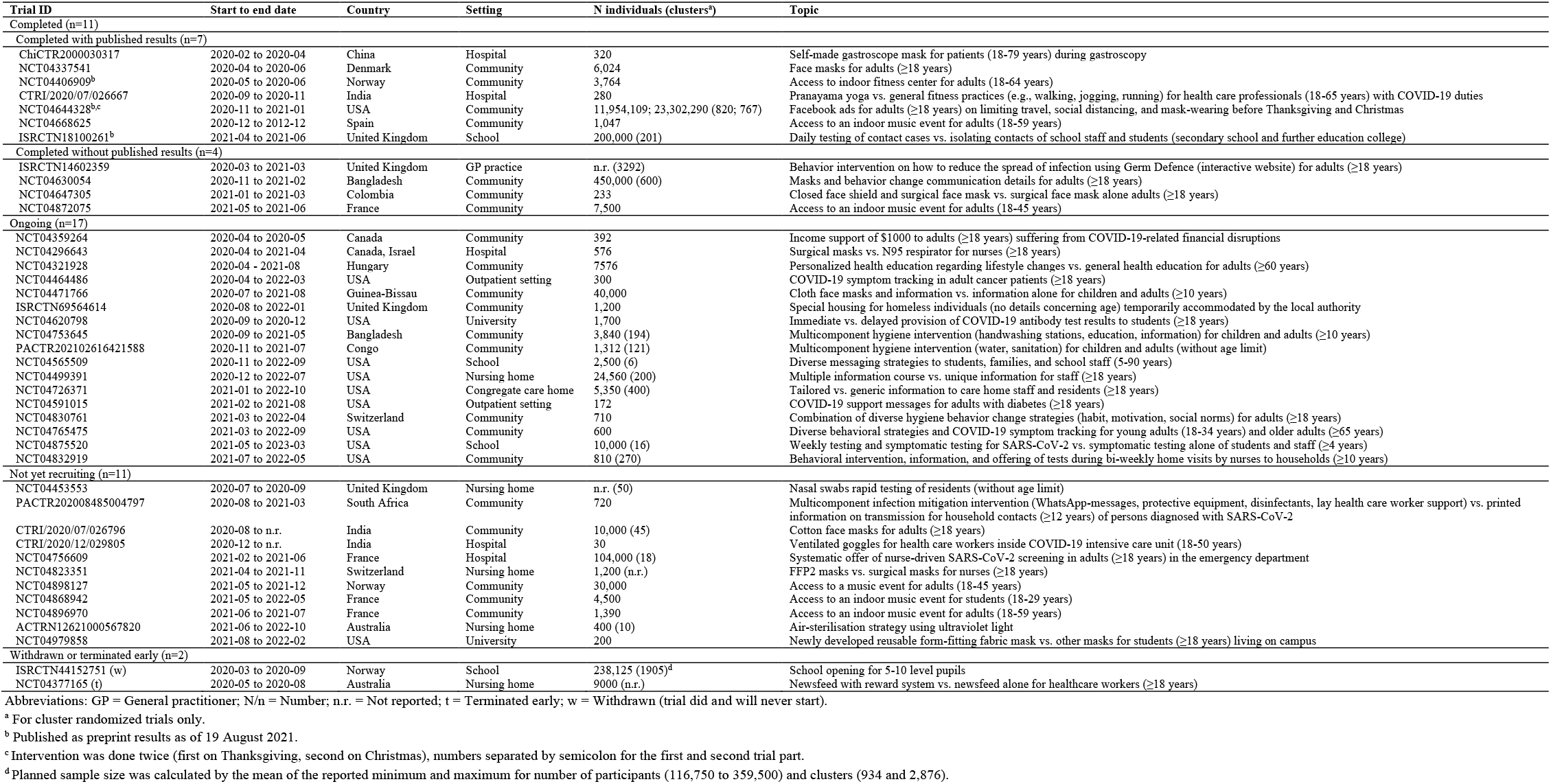
Characteristics of all 41 randomized trials on non-pharmaceutical interventions to prevent COVID-19 as of August 2021

The types of NPIs were diverse. Of the 41 trials, 11 trials (26.8%) assessed protective equipment such as various masks, face shields or goggles; 9 trials (22.0%) assessed diverse COVID-19-related information and education programs (e.g., text messages, newsfeeds, personalized health education, Facebook advertisements, nurse-led home visits); 5 trials (12.2%) assessed the impact of attending a musical mass event/concert under specific safety measures; 5 trials (12.2%) assessed diverse testing and screening strategies in nursing homes, schools, universities, and the emergency department; 5 trials (12.2%) assessed multicomponent behavioral interventions on hygiene management in the community. The other 6 trials assessed access to a fitness center, school opening, special housing of homeless individuals, direct income support for individuals with COVID-19-related financial disruptions, yoga sessions for health care professionals, and COVID-19-related symptom tracking in outpatients with cancer.

The trials planned to recruit a median of 1,700 participants (IQR, 588 to 9,500, range 30 to 35,256,399) with a median planned duration of 8 months (IQR, 3 to 14, range 1 to 24). The largest trial assessed the impact of Facebook advertisements to prevent traveling and family reunions during Thanksgiving and Christmas with >35 million participants ^12^, the shortest trial assessed the audience infection rate 8 days after an indoor music event.^13^ A typical randomized trial on NPIs randomly allocated individuals (n=23, 56.1%), rather than clusters (n=18, 43.9%), to two comparison groups (n=38, 92.7%) and assessed a COVID-19-related primary outcome (n=33, 80.5%) that was laboratory-confirmed (n=33, 80.5%).

## Discussion

Worldwide, 41 randomized trials assessing NPIs have been initiated with published results available to inform policy decisions for only 7 of them. This is a tiny fraction of more than 4000 randomized trials on COVID-19 registered globally as of August 2021.^11^ Given the unprecedented public health impact worldwide and the wide debates about benefits and harms of NPIs, this number seems disproportionate in light of other interventions that have been the focus of much more evaluation with randomized trials. For example, over 300 randomized trials included the highly debated hydroxychloroquine in their intervention arm^14^ which has been found to be associated with increased mortality in COVID-19 patients. ^15^

The list of available treatments for COVID-19 is limited ^16,17^ and with vaccination rates plateauing in many countries ^18^, NPIs and their effectiveness will remain a central debate. Our analysis clearly indicates that randomized trials are generally feasible to assess the benefits and harms of NPIs during a pandemic situation, in a very short time using the most reliable method to provide reliable evidence for optimal health policy decision making. The World Health Organization (WHO) is committed to improving the evidence base on effectiveness of public health and social measures^19^, and this overview may inform further developments in this important area of evidence generation.

This study is limited as our sample relies on accurate trial registration and reporting. We cannot exclude that some NPI randomized trials might have been missed because they were not registered, outcomes related to SARS-CoV-2-infections were incorrectly or not reported in the registries, or because of our single-reviewer screening approach. However, it is unlikely that we have been unaware of pertinent results of further NPI trials, given their substantial impact on current debates and scarcity of the evidence.

## Conclusions

Overall, during the first 18 months of the pandemic, the worldwide clinical research agenda failed to provide urgently needed evidence determining best strategies to prevent COVID-19 in schools, workplaces, nursing homes and other settings substantially affected. A long-term research agenda including behavioral, environmental, social, and systems level interventions is urgently needed to guide policies and practices in the current and future public health emergencies.

## Supporting information

PRISMA checklist

## Data Availability

All data generated or analyzed during this study are included in this publication.

## Notes

Funding: The COVID-evidence project is supported by the Swiss National Science Foundation, project ID 31CA30_196190.

### Competing Interest Statement

The authors have declared no competing interest.

### Funding Statement

The COVID-evidence project is supported by the Swiss National Science Foundation, project ID 31CA30_196190.

### Author Declarations

Ethical approval is not necessary for this study.

## References

1. World Health Organization. Overview of public health and social measures in the context of COVID-19: Interim guidance. Accessed July 25, 2021. https://www.who.int/publications/i/item/overview-of-public-health-and-social-measures-in-the-context-of-covid-19

2. Doroshenko A. The Combined Effect of Vaccination and Nonpharmaceutical Public Health Interventions-Ending the COVID-19 Pandemic. JAMA Netw Open. 2021;4(6):e2111675. doi:10.1001/jamanetworkopen.2021.11675

3. Cristea IA, Naudet F, Ioannidis JPA. Preserving equipoise and performing randomised trials for COVID-19 social distancing interventions. Epidemiol Psychiatr Sci. 2020;29:e184. doi:10.1017/S2045796020000992

4. Fretheim A, Flatø M, Steens A, et al. COVID-19: we need randomised trials of school closures. J Epidemiol Community Health. 2020;74(12):1078–1079. doi:10.1136/jech-2020-214262

5. Ioannidis JPA. A fiasco in the making? As the coronavirus pandemic takes hold, we are making decisions without reliable data. STAT. Published 2020. Accessed August 19, 2021. https://www.statnews.com/2020/03/17/a-fiasco-in-the-making-as-the-coronavirus-pandemic-takes-hold-we-are-making-decisions-without-reliable-data/.

6. Janiaud P, Axfors C, van’t Hooft J, et al. The worldwide clinical trial research response to the COVID-19 pandemic - the first 100 days [version 2; peer review: 2 approved]. F1000Res. 2020;9:1193. doi:10.12688/f1000research.26707.2

7. Burns J, Movsisyan A, Stratil JM, et al. International travel-related control measures to contain the COVID-19 pandemic: a rapid review. Cochrane Database Syst Rev. 2021;3:CD013717. doi:10.1002/14651858.CD013717.pub2

8. Nussbaumer-Streit B, Mayr V, Dobrescu AI, et al. Quarantine alone or in combination with other public health measures to control COVID-19: a rapid review. Cochrane Database Syst Rev. 2020;9:CD013574. doi:10.1002/14651858.CD013574.pub2

9. Regmi K, Lwin CM. Impact of non-pharmaceutical interventions for reducing transmission of COVID-19: a systematic review and meta-analysis protocol. BMJ Open. 2020;10(10):e041383. doi:10.1136/bmjopen-2020-041383

10. Page MJ, McKenzie JE, Bossuyt PM, et al. The PRISMA 2020 statement: an updated guideline for reporting systematic reviews. BMJ. 2021;372:n71. doi:10.1136/bmj.n71

11. COVID-evidence Database. Planned, ongoing and completed trials to treat and prevent COVID-19. Published July 27, 2021. Accessed July 27, 2021. https://covid-evidence.org/database

12. Breza E, Stanford FC, Alsan M, et al. Doctors’ and Nurses’ Social Media Ads Reduced Holiday Travel and COVID-19 infections: A cluster randomized controlled trial in 13 States: Preprint;2021. doi:10.1101/2021.06.23.21259402

13. Revollo B, Blanco I, Soler P, et al. Same-day SARS-CoV-2 antigen test screening in an indoor mass-gathering live music event: a randomised controlled trial. The Lancet Infectious Diseases. 2021;Article in Press. doi:10.1016/S1473-3099(21)00268-1

14. Janiaud P, Hemkens LG, Ioannidis JP. Challenges and lessons learned from Covid-19 trials – should we be doing clinical trials differently? Canadian Journal of Cardiology. 2021;Article in Press. doi:10.1016/j.cjca.2021.05.009

15. Axfors C, Schmitt AM, Janiaud P, et al. Mortality outcomes with hydroxychloroquine and chloroquine in COVID-19 from an international collaborative meta-analysis of randomized trials. Nat Commun. 2021;12:2349. doi:10.1038/s41467-021-22446-z

16. U.S. Food and Drug Administration. Emergency Use Authorization: Coronavirus Disease 2019 (COVID-19) EUA Information. Accessed August 19, 2021. https://www.fda.gov/emergency-preparedness-and-response/mcm-legal-regulatory-and-policy-framework/emergency-use-authorization#covid19euas

17. European Medicines Agency. Treatments and vaccines for COVID-19. Published August 19, 2021. Accessed August 19, 2021. https://www.ema.europa.eu/en/human-regulatory/overview/public-health-threats/coronavirus-disease-covid-19/treatments-vaccines-covid-19

18. Blamont M, Erman M, Lubell M. Vaccination burnout?: Delta variant spurs countries to speed up shots. Published 2021. Accessed August 19, 2021. https://graphics.reuters.com/HEALTH-CORONAVIRUS/VACCINATION/xklvyxrdgpg/.

19. World Health Organization. Director-General’s opening remarks at the media briefing on COVID-19 – 14 June 2021. Published August 4, 2021. Accessed August 17, 2021. https://www.who.int/director-general/speeches/detail/director-general-s-opening-remarks-at-the-media-briefing-on-covid-19-14-june-2021

20. Janiaud P, Axfors C, Saccilotto R, Schmitt AM, Hirt J, Hemkens LG. COVID-evidence: a living database of trials on interventions for COVID-19. Accessed May 31, 2021. https://osf.io/gehfx/

